# Innovative mHealth Intervention Providing Sustained Anticipatory Guidance (Zero Cavity): Design, Validation, User Perception and Effectiveness – A Randomized Controlled Trial Protocol

**DOI:** 10.64898/2026.09.11.26362881

**Authors:** MS Muthu, Vineet Dhar, Ankita Saikia, S Vandana, R Harini, Kalpana Balakrishnan, Latha Nirmal, P Umapathy, John Davis

## Abstract

**Introduction:** Early Childhood Caries (ECC) remains one of the most common chronic conditions in childhood, affecting over 600 million children globally. When untreated, it can lead to severe pain, abscesses, difficulty in chewing that contributes to malnutrition, impaired speech, disrupted sleep, low self-esteem, social withdrawal, and poor academic performance, all of which diminish a child’s overall quality of life. Although largely preventable, ECC remains highly prevalent because of delayed preventive care and low parental awareness. Effective health education needs to be consistent, accessible, and available whenever parents need it. Although current evidence indicates that anticipatory guidance is a proven approach, its long-term reinforcement through a mobile app has not yet been tested. This study aims to develop and evaluate the effectiveness of a mobile health (mHealth) application delivering Sustained Anticipatory Guidance (SAG) for the prevention of ECC.

**Methods and analysis:** A two-arm parallel hospital-based randomized controlled trial will be conducted at Sri Ramachandra Institute of Higher Education & Research, Chennai, India. The study will enrol 540 mother-infant dyads from the departments of Pediatric Medicine and the Centre for Early Childhood Caries Research (CECCRe). Eligible dyads with infants aged 6–12 months in the predentate stage will be randomized to the “Zero-Cavity” mHealth app (intervention) or standard care (control). Baseline and semi-annual data on oral hygiene, fluoride exposure, and feeding/diet will be collected. Calibrated dentists will perform quarterly clinical exams using ICDAS II criteria, recording cavitated and non-cavitated lesions. For children at high risk or with enamel defects/incipient lesions, clinical photographs will be taken to support individualized preventive guidance. The primary outcome will be the effectiveness of the app in preventing ECC, while the secondary outcomes will include ECC incidence and prevalence.

**Discussion:** Despite being largely preventable, ECC remains a major public health challenge, sustained by limited preventive access and inconsistent counselling, especially in LMICs. This trial evaluates the Zero Cavity app, believed to bridge this gap by providing sustained, age-appropriate anticipatory guidance beyond clinic settings on oral development, hygiene, diet/feeding, and fluoride use. The behavioural theory-driven approach is expected to enhance cultural relevance, usability, and behaviour change, with optimism that process evaluation will illuminate engagement and implementation. It is believed that this large, adequately powered, parallel-group RCT with intention-to-treat analysis will offer, with optimism, a scalable, low-cost model that will expand the reach and consistency of continuous preventive care in diverse urban and peri-urban populations.

**Trial registration:** CTRI/2026/04/107305

## Introduction

Early Childhood Caries (ECC) is among the most common chronic diseases of childhood, affecting more than 600 million children worldwide [1]. The prevalence of ECC in low- and middle-income countries (LMIC) ranges from 30%-52%, and children born into low-income families are particularly vulnerable [2]. If left untreated, this disease can cause significant pain, abscesses, chewing difficulty leading to malnutrition, poor speech articulation, poor sleep habits, low self-esteem, social withdrawal, and poor school performance that leads to a diminished overall quality of life [3,4]. Literature on ECC prevention and management highlight’s early introduction and sustained interventions as the best care method to prevent the disease [5,6].

Despite being largely preventable, ECC continues to affect a substantial proportion of children due to delayed preventive care and limited parental awareness. Anticipatory guidance (AG) delivered early in life is effective in preventing ECC, but its reach is often inconsistent in routine clinical practice. Although the current Indian health system highlights growing ECC concerns, critical gaps limit access to and utilisation of care. In recent years, digital health has emerged as a promising means of disseminating routine and innovative information to address health needs [7]. With the substantial increase in smartphone use, app-based health education has become feasible, and mobile health (mHealth) interventions now offer an opportunity to deliver standardized, timely, and scalable preventive guidance. Prior studies [8–12] have shown that digital health interventions can improve health knowledge and behaviours when grounded in behavioural theory. Evidence-based oral health interventions delivered to parents and children via mobile phones have been shown to improve adherence to oral hygiene practices. This behavioural change has been linked to the Health Belief Model [13], the Transtheoretical Model [14], or the Theory of Planned Behaviour [15]. Research has shown that Motivational Interviewing (MI) and caries prevention activities often increase knowledge but do not necessarily improve oral health behaviours or reduce caries incidence [16,17].

Health information should be repetitive and available at all points in time whenever there is a need [18]. However, anticipatory guidance, followed through mobile app-assisted sustenance, has not been tested so far. We hypothesize that an easily accessible and user-friendly mobile application can serve as an effective oral health intervention. Therefore, in collaboration with professional app developers, we will develop the Zero Cavity mobile app to provide continuous, age-appropriate guidance. We will evaluate its effectiveness in a randomized trial of 540 mother-child dyads in Chennai, Tamil Nadu, India. Using an intent-to-treat analysis, we will compare SAG delivered via the app with traditional anticipatory guidance provided at routine dental visits for improving oral health outcomes in this diverse LMIC population.

## Materials and Methods

### Ethical Approval, Permissions, and Informed Consent

The study was approved by the Institutional Ethics Committee of Sri Ramachandra Institute of Higher Education and Research, Chennai [IEC No: NI/23/APR/86/06] and registered in Clinical Trials Registry - India [CTRI/2026/04/107305]. Permission will be sought from the heads of Pediatric Medicine and the Centre for Early Childhood Caries Research (CECCRe) to recruit the participants. Voluntarily signed informed consent will be obtained by the project coordinator from the parent/primary caregiver for study participation, collection of data, clinical oral examination, and recording of intraoral photographs. The participants will be informed about the study objectives, procedures, potential risks and benefits, and their right to withdraw at any time. Findings will be reported in accordance with Standard Protocol Items: Recommendations for Interventional Trials (SPIRIT) 2025 guidelines [19] for randomized controlled trials. The completed SPIRIT checklist that outlines the location of each item is provided as Supporting Information (S1 checklist).

### Study design and study setting

This study will utilize a parallel-group, two-arm randomized controlled trial design conducted at Sri Ramachandra Institute of Higher Education & Research, a tertiary care teaching hospital in Chennai, India. The Pediatric Medicine and the Centre for Early Childhood Caries Research (CECCRe) outpatient departments of the hospital serve a diverse urban and peri-urban population and will function as the primary site for participant recruitment.

### Sample size

The sample size calculation was based on a previous study [6] reporting ECC prevalence as 40%. Study power was set at 80% to detect a 15% difference between group proportions. This was tested using a two-sided Z-test with a pooled variance test statistic. Based on these parameters, 244 participants per arm were required. The significance level was set at 0.05. With an estimated attrition rate of 10%, a total of 270 participants per arm will be enrolled, yielding an overall sample size of N = 540.

### Patient and public involvement

Patients and the public were not directly involved in the design, recruitment, or conduct of this study.

### Eligibility criteria

Parents or primary caregivers of infants aged 6–12 months who can speak, read and write English will be eligible to participate in the study. Participants must be willing to provide written informed consent and to attend all scheduled follow-up visits during the study period. Infants with systemic conditions/syndromes or developmental disorders will be excluded.

### Recruitment

Participant recruitment commenced in April 2026 and is anticipated to be completed by December 2026. The recruitment team will comprise a project coordinator and two trained dental professionals (a pediatric dentist and an oral pathologist). Recruitment will be through in-person meet-and-greet by trained staff using visual aids (MAAC chart/anticipatory guidance), referral of pre-dentate infants by paediatricians, and display of printed/Tamil-English flyers, mobile standees, e-flyers via WhatsApp, and animated oral-health videos with contact number on television screens in Pediatric Medicine OPD and CECCRe waiting areas. Either of the trained dentists will approach eligible parents in the OPD and provide a brief explanation of the study and its eligibility criteria. Interested and eligible participants will be escorted to a private room where the study will be explained, and the project coordinator will obtain informed consent in English. Consent forms will include a study description, respondent burden, potential risks and benefits, and names and contact information of individuals who can be contacted for additional study details. During the consent process, the project coordinator will clearly explain to the participants that their child’s participation in the study is voluntary and that their decision to participate or not will not in any way affect their care at the hospital. All participants’ parents/caretakers will receive a copy of the consent form for their records. No potential participants will be excluded based on gender, race, caste or ethnicity. Each study participant will receive a personal ID number. To protect confidentiality, the link between personal identifiers and study information will be coded, and only the investigators will have access to it. The research team will facilitate the baseline data collection, pediatric dental appointments, and subsequent follow-up visits.

### Randomization and allocation concealment

The participants will be divided into 2 groups as follows: Group 1 - Anticipatory Guidance (AG) at initial and 6-monthly routine visits (n=270); Group 2 - Anticipatory Guidance (AG) at initial and 6-monthly routine visits plus SAG through mobile App (n=270). Participants will be randomized to one of the two arms in a 1:1 ratio using a computer-generated list with stratification. The allocation sequence will be generated by a surveyor not otherwise involved in the trial, using variable length random permuted blocks of sizes 4 and 6. Sequentially numbered, opaque, sealed envelopes containing allocation sequences will be prepared before the trial. A researcher not directly involved in the selection process will conduct the randomization and assignment of participants to the groups. The trained dentists will carry out the examinations at baseline and recalls.

### Training and Calibration

The trained dentists involved in participant recruitment will undergo training in ICDAS II (International Caries Detection and Assessment System) criteria to diagnose dental caries both clinically and radiographically. Training will be completed through the e-learning module available on the ICCMS website. This will be followed by a 2-day calibration exercise conducted in the Department of Pediatric Dentistry. During calibration, both examiners will independently examine three children attending the outpatient clinic each day. Findings from the two examiners will be compared to identify discrepancies in interpretation. Examinations will be repeated until satisfactory inter-examiner agreement is achieved. For reliability assessment, both examiners will independently examine three children aged 6 months to 3 years with carious lesions corresponding to ICDAS II codes over two days. An experienced investigator (MSM, Principal Investigator) will supervise the process. Inter- and intra-examiner reliability will be assessed by comparing scores between examiners, and Cohen’s kappa coefficients will be calculated.

### Interventions

Recruited mothers of participants in both Group 1 and Group 2 will receive comprehensive Anticipatory Guidance (AG) on infant oral health, delivered one-on-one using audiovisual aids. Demonstrations of oral hygiene measures will also be provided.

Group 1 participants will receive AG only.

Group 2 participants will receive AG plus SAG delivered through a mobile application. Mothers in Group 2 will receive a smartphone with the ’Zero Cavity’ app pre-installed and an included data plan. They will be encouraged to follow SAG delivered via weekly and monthly age-appropriate notifications covering the four domains of anticipatory guidance: oral development, fluoride adequacy, oral hygiene, and nutrition and diet. The design and flow of the randomized controlled trial are summarized in Fig 1.

**Fig 1.**
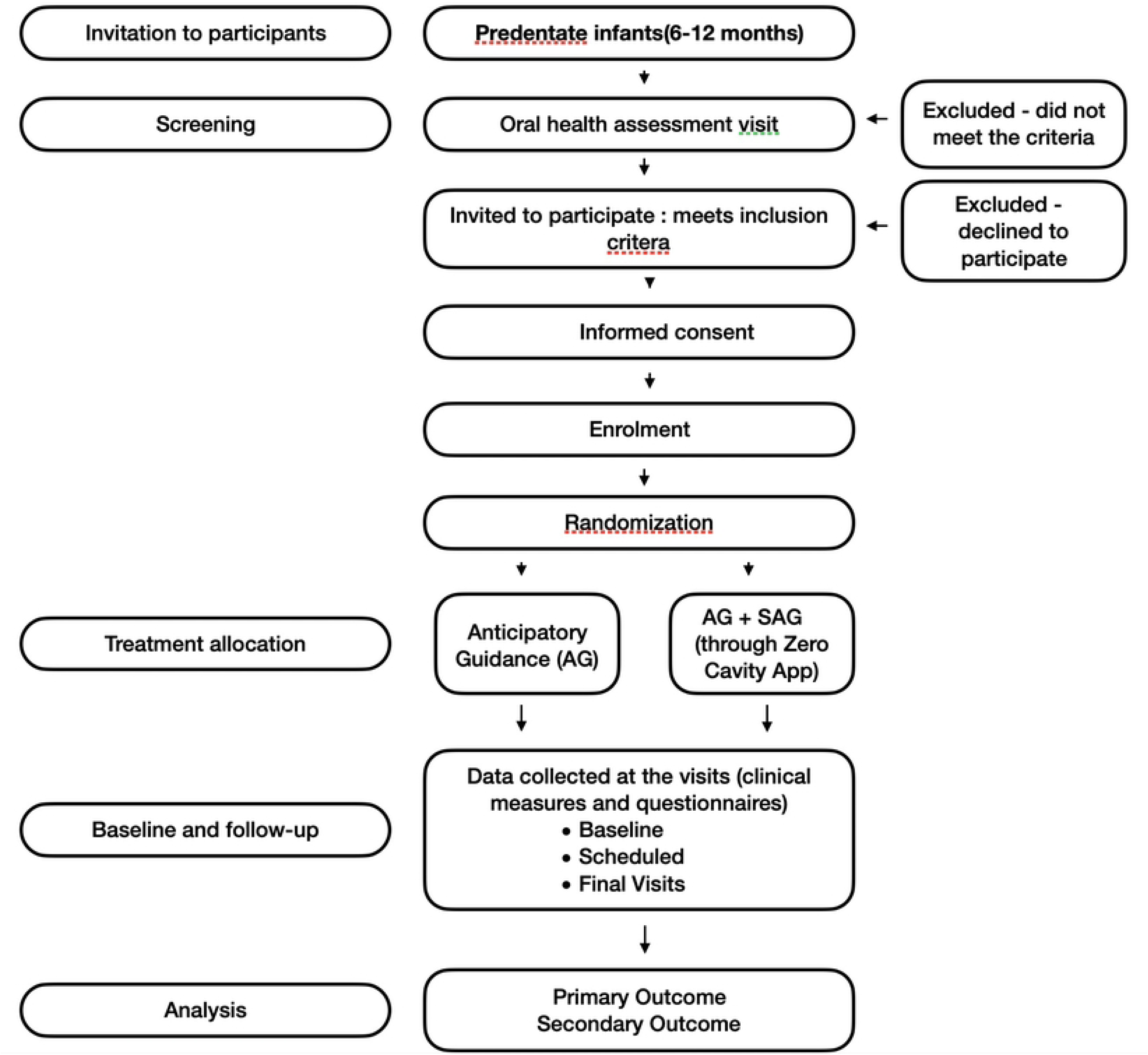
Flowchart of Study Design and Participant Flow in the Two-Arm Randomized Controlled Trial. Legend: AG, Anticipatory Guidance; SAG, Sustained Anticipatory Guidance

### Primary and Secondary Outcomes

The overall effectiveness of the app will be assessed by criteria reported by Abirami et al.^19^ as the primary outcome. The intervention will be regarded as Highly Effective (Cavitated and Non-cavitated lesion = 0), Moderately Effective (Cavitated lesion = 1-2; non-cavitated lesion = 1-4), Least effective (Cavitated lesion = greater than or equal to 3; non-cavitated lesion greater than or equal to 5), as described. Occurrence of ECC will be reported as incidence and prevalence as secondary outcomes.

### Data collection

#### Baseline data

All collected baseline data will be linked to a study participant ID number. Questionnaires will be collected at enrolment to assess socio-demographic status, oral hygiene measures, fluoride history, awareness of ECC and Oral health, feeding and diet histories. The research team will be responsible for data collection at the departments of paediatrics and pediatric dentistry using widely accepted methods for interviewing and dental caries data collection. All questionnaires will be adapted to similar previous studies that have been field-tested, and then modified to best suit the proposed study population.

#### Follow-up data

Data on oral hygiene measures, fluoride history, feeding and diet histories will be collected semi-annually. The infants will be examined quarterly in a knee-to-knee position, under illumination, using ICDAS II criteria by either of the calibrated dentists. The teeth will be initially assessed wet and then air-dried. Cotton rolls will be used to clean the teeth of food debris and to dry them. The examiners using the “lift the lip technique” will assess the labial, palatal, mesial, and distal surfaces of each tooth and record the findings in a form designed for the study. Dental caries will be scored as cavitated and non-cavitated lesions. For children at high risk and showing enamel defects/hypoplasia/incipient lesions, a clinical photograph of the tooth will be taken to promote oral health education and individualise preventive AG protocols. After scoring the teeth, the camera lens will be positioned parallel to the tooth surface. The shutter speed and camera (Shofu Eye Special IV) aperture will be set and recorded. Labial tooth surfaces will be imaged under both flash and strip light illumination. Each image will be checked for quality and re-taken if necessary. The images will be saved as tagged image format files (TIFFs). Any child showing cavitated lesions or needing dental care for other reasons will be referred for further treatment to the Department of Pediatric Dentistry.

### Data safety and management

Participant information will be de-identified and coded, and all study data will be stored securely in password-protected files and locked cabinets accessible only to authorised study personnel. All study investigators will undergo training in Human Subjects Research Ethics. Study participants will be assured that information gathered during interviews and health assessments will be kept confidential. A Data Safety and Monitoring Board (DSMB) will be established to oversee the safety and efficacy of the full RCT. The team will comprise internal and external members who will meet biannually in online or offline mode to review trial progress and data safety, and provide recommendations regarding study status.

The interventions are behavioral and educational with low anticipated risk. All adverse events including participant distress, minor issues during dental examination, unexpected clinical findings, or data privacy concerns will be documented and reported within 24 hours of awareness, with data handled using coded identifiers to maintain confidentiality. Any unanticipated problems involving risk to participants will be reported to the DSMB, Institutional Ethics Committee and NIH per applicable guidelines. If a substantial reduction in cavities demonstrates clear benefit, the DSMB may determine that clinical equipoise no longer exists and recommend early termination to allow control arm participants to benefit from the intervention.

### Data analysis

Efficacy Population: For the efficacy analysis, an intention-to-treat (ITT) and modified intention-to-treat (mITT) approach will be adopted, with the ITT analysis being the primary approach, to provide a comparison of the different groups.

Intention to Treat: To provide a pragmatic comparison of the different groups, the principle of intention-to-treat will be the main strategy of analysis adopted for the primary and secondary endpoints. These analyses will be conducted on all patients assigned to the treatment groups as randomized, regardless of the treatment received.

Modified Intention to Treat: A modified intention-to-treat approach will be used for secondary analyses. Patients with protocol violations will be censored from the respective episode onwards.

### Statistical methods

Demographics and baseline characteristics will be summarized for all screened patients, those who meet the study inclusion criteria, those who are eligible and randomized, those who are eligible but not randomized, those who withdraw from the study after randomization, and those who are lost to follow-up. These data will be presented in CONSORT flow diagrams. The number of patients discontinuing from the study will be tabulated by reason for study discontinuation. The number (%) of patients attending the scheduled follow-up will be reported. The baseline value is defined as the last available value before randomization. For continuous variables, descriptive statistics including n, mean, standard deviation, minimum, 1st quartile, median, 3rd quartile, and maximum will be calculated. For categorical variables, counts and percentages will be provided. All statistical tables will include p-values, which will be reported to at least three decimal places as generated by the statistical software. P-values less than 0.001 will be reported as “<0.001” based on the current version of SPSS. Unless stated otherwise, all statistical tests will be two-sided with a significance level set at α = 0.05. Results of the study are anticipated by May 2028.

### Analysis of Primary and Secondary Outcomes

Overall Effectiveness: The effectiveness of the Zero cavity app will be assessed at the end of the study period using 3-point ordinal data. Therefore, we will be using the Mann-Whitney U test for between-group comparisons.

Prevalence of ECC: At the end of the study, prevalence of ECC will be reported as the absolute number (%) of cavitated and non-cavitated lesions in both groups. Surface-wise caries occurrence will be assessed using ICDAS scores and reported as mean values. The chi-square test will be applied to compare the between-group percentage (prevalence of ECC) estimates. ANCOVA test will be employed to compare the mean ICDAS scores between the two groups. To compare the data within each group at different time intervals, we will use a chi-square test and repeated measures ANOVA for categorical and continuous data, respectively.

Incidence of ECC: Incidence of ECC at the last follow-up will be measured as Incidence Density. Incidence density for risk of caries of a person (IDp) and a tooth surface (IDs) will be summarized as follows

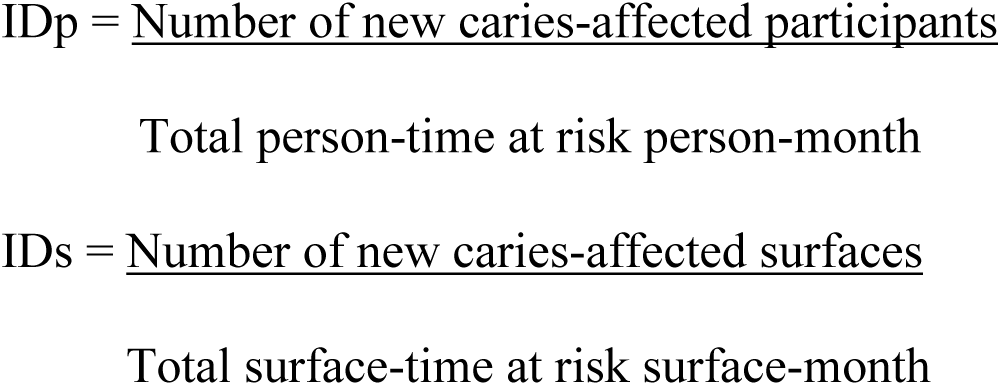

Transitional Status of each tooth and the surface (Transitional Probability and Markov Model): Transitional probability will be calculated by measuring the transitional changes in the dental status. This will be based on the tooth-wise patterns from baseline to the first follow-up and in the subsequent follow-ups until the end of the study. To calculate the transition probabilities, we will use the maximum likelihood method. That is, each transition probability was estimated by the number of observed transitions during a given cycle, divided by the total number of all possible observations during that cycle.

Analysis of dietary and oral hygiene practices: Descriptive statistics will be used to estimate the frequencies and mean (SD) of study variables. Unpaired t-test and Chi-square test will be used to compare the continuous data and categorical data between the groups. Pearson chi-square test will be used to assess the association of study risk factors with the outcome of ECC (Yes/No). Bivariate and Multivariate logistic regression analysis will be performed to estimate the unadjusted and adjusted odds ratio with corresponding 95% confidence intervals (CI), respectively. The confounders which shows the association in bivariate analysis with P ≤ 0.10 will be controlled to estimate the adjusted Odds Ratio.

The level of statistical significance in the present study will be set at p ≤ 0.05 for all tests.

### Missing Values

Efficacy: For participants with incomplete data (for dropping out or for any other reason), missing values will not be imputed.

Safety: No imputation will be done on missing safety data, unless otherwise stated.

## Discussion

ECC continues to pose a major public health challenge, even though it is largely preventable. Persistent barriers such as limited access to timely preventive services, inconsistent parent counselling, and inadequate reinforcement of oral health practices sustain a high disease burden, especially in low- and middle-income settings. This study aims to address these gaps by testing a mobile health–based anticipatory guidance intervention for parents of young children. By leveraging digital tools to deliver standardized oral health education and support, the intervention seeks to improve both the reach and consistency of preventive care during early childhood.

The Zero Cavity mobile application aims to provide sustained, age-appropriate anticipatory guidance that extends beyond the constraints of routine healthcare settings. By delivering standardised information across key domains - oral development, oral hygiene practices, diet and feeding behaviours, and fluoride use- the intervention seeks to promote consistent preventive behaviours during critical periods of tooth eruption and development. The use of behavioural theory to guide content development strengthens the potential for meaningful behaviour change. This digital approach enhances cultural relevance, usability, and acceptability of the intervention while allowing rigorous evaluation of effectiveness. The inclusion of a process evaluation will provide insight into participant experiences, implementation fidelity, and contextual factors influencing engagement, which are essential for interpreting trial outcomes.

The primary strength of this study is its large, adequately powered, parallel-group randomized controlled trial design with an intention-to-treat analysis, which enhances the validity and generalizability of the findings. Furthermore, it evaluates a scalable, low-cost mobile app-based intervention for delivering continuous, age-appropriate anticipatory guidance in a low- and middle-income country (LMIC) setting, addressing a critical gap in preventive care between clinic visits among a diverse urban and peri-urban population. However, the study has certain limitations. The requirement of smartphone access may limit participation and affect equity, and the nature of the behavioural intervention precludes blinding of participants and personnel, which may introduce bias.

The findings of this study will be disseminated through publications in peer-reviewed journals and presentations at scientific conferences, regardless of the outcome. De-identified participant data will be made available upon reasonable request to the corresponding author after publication of the main results.

## Author contributions

MSM conceived the study. MSM, VD, and AS designed the methodology. HR and VS drafted the original manuscript. MSM, AS, VD, KB, UP, LN, JD critically revised the manuscript for important intellectual content. All authors reviewed and approved the final version and agree to be accountable for all aspects of the work. MSM is the guarantor.

## Acknowledgements

We would like to thank Dr Selva Arockiam for the statistical assistance. We would also like to acknowledge iThenticate (http://www.ithenticate.com) for checking plagiarism.

## Supporting information

The SPIRIT 2025 checklist for this study protocol is provided as Supporting Information (S1 Checklist)

**S1 checklist. SPIRIT 2025 Checklist for Zero Cavity Trial Protocol.**

## Data availability

The data that support the findings of this study are available from the corresponding author upon reasonable request.

